# Longitudinal Associations of Chemotherapy-Induced Symptom Clusters with Healthcare Utilization and Mortality among Patients with Gastrointestinal Cancers

**DOI:** 10.1101/2025.10.08.25337569

**Authors:** Chengbo Zeng, Nneka N. Ufere, Patricia C. Dykes, Shumenghui Zhai, Yu-Jen Chen, Jiancheng Ye, Maria O. Edelen, Andrea L. Pusic, Jason B. Liu, Kelsey S. Lau-Min, Kelly M. Kenzik

**Author notes:** **Corresponding author:** Chengbo Zeng, Ph.D., Department of Surgery, Brigham and Women’s, Hospital Address: 1620 Tremont Street, Boston, MA 02120. These authors contributed equally to this work (Jason B. Liu, Kelsey S. Lau-Min, Kelly M. Kenzik). **Author Contributions:** Conceptualization (CZ, NNU, JBL, KSL, KMK), Data curation (CZ, YJC, ALP), Formal analysis (CZ, YJC), Investigation (All authors), Methodology (CZ, YJC, JY, MOE, KMK), Project administration (CZ, YJC), Resources (ALP), Software (CZ, YJC), Supervision (JBL, KSL, KMK), Validation (SZ, PCD, NNU, KSL, JBL), Visualization (CZ, YJC), Writing – original draft (CZ), Writing – review & editing (All authors).

## Abstract

**Background:** Chemotherapy (CTX) induced neurological and gastrointestinal (GI) symptom clusters are common, but their longitudinal associations with healthcare utilization and mortality remain unclear.

**Patients and Methods:** We conducted a retrospective study of 973 GI cancer patients in the Mass General Brigham Health System from 2019 to 2024. Patient-reported symptoms, including constipation, decreased appetite, diarrhea, dyspnea, fatigue, fever, insomnia, nausea, paresthesia, pain, rash, and vomiting, were routinely collected at CTX initiation, days 30, 60, or 90. Primary outcomes were all-cause urgent care visits, emergency department (ED) visits, and death within one year of CTX. Neurological cluster was defined by the presence of fatigue, insomnia, paresthesia, and pain. GI cluster included constipation, decreased appetite, diarrhea, nausea, and vomiting. Time-to-event analysis was used to predict each outcome based on changes in neurological and GI clusters over the first 90 days of CTX. We also examined whether findings varied by age group, comorbidity level, time since diagnosis, cancer stage, and type.

**Results:** Over time, the burden of neurological symptoms was significantly higher than that of GI symptoms. Higher neurological burden was associated with an increased risk of urgent care visits (Adjusted HR: 1.27 [95% CI: 0.97–1.67]), but not ED visits or death, with stronger associations among older adults and those without comorbidities. In contrast, higher GI burden was associated with greater risks of ED visits (Adjusted HR: 1.10 [95% CI: 1.02–1.19]) and death (Adjusted HR: 1.08 [95% CI: 0.94–1.24]), but not urgent care visits, with stronger effects among older adults, patients with comorbidities, and those with advanced cancers.

**Conclusions:** Neurological and GI clusters were common among patients with GI cancers receiving CTX. Their short-term changes exhibited differential associations with long-term healthcare utilization and mortality.

## Background

Chemotherapy (CTX), a central component of treatment for gastrointestinal (GI) cancers can lead to severe symptoms, which contribute to increased healthcare utilization and higher mortality. ^1,2–8^ Existing studies have examined the associations between individual symptoms and overall symptom burden and these outcomes.^9–11^ Yet, symptoms often co-occur and form distinct clusters, such as “nausea-vomiting” and “fatigue-insomnia.”^12–15^ A symptom cluster may share a common underlying mechanism or arise from a primary symptom that triggers secondary ones.^12,16–18^ As a result, different symptom clusters may have varying associations with clinical outcomes.

Neurological and GI symptoms are two major CTX-induced clusters.^5,8,19^ Commonly administered CTX agents for GI cancers, such as 5-FU, oxaliplatin, irinotecan, and paclitaxel, cause significant toxicities affecting the nervous and digestive systems.^11,20–23^ These agents form the backbone of widely used regimens for GI cancers, including FLOT, FOLFOX, and FOLFIRI, which may amplify the toxicities of individual agents and result in a broad spectrum of symptoms involving both neurological damage and GI side effects.^9–11,20–32^ Individual symptoms within a cluster may change during CTX, leading to varying burdens of neurological and GI clusters. Currently, their longitudinal associations with healthcare utilization and mortality remain unclear. Evidence is lacking on whether their short-term changes are associated with long-term healthcare utilization and mortality, and which cluster has a stronger impact. This knowledge, if found, could help clinicians to prioritize symptom management, alleviate adverse side effects, and ultimately improve cancer care.

We sought to analyze the burden of GI and neurological clusters over time and examine their associations with one-year healthcare utilization and mortality among patients receiving CTX for GI cancers. Specifically, we aimed to address three research questions: (1) What are the most prevalent co-occurring symptoms within each cluster during the first 90 days of CTX? (2) Are the short-term changes in the burden of these two clusters associated with the risks of one-year healthcare utilization and mortality? and (3) Are these associations, if present, consistent across prognostic groups?

## Methods

### Study design, setting, and data resource

This was a retrospective cohort study using electronic medical records (EHR) and patient-reported symptom data routinely collected during the care of GI cancer patients within the Mass General Brigham (MGB) Health System. Symptom assessment is part of the MGB PROMs Program, details of which had been described in prior research.^33–36^ The Patient-Reported Outcome (PRO) version of the Common Terminology Criteria for Adverse Events (PRO-CTCAE^®^) was used to assess symptoms. Patients with a scheduled appointment were asked to complete symptom assessments one week before their visit, using either the EHR’s patient portal or on tablets provided at the clinic for those who did not complete them beforehand. The reported scores were made available to clinicians and patients to support treatment planning and clinical decision-making.

### Study population

Between 01/2019 and 01/2024, we identified adult patients who met the following criteria: (1) had a diagnosis of all-stage colorectal, esophageal, hepatobiliary, pancreatic, or gastric cancers based on ICD-10 codes**(Supplemental Table S1)**, (2) initiated any cytotoxic chemotherapy to GI cancers, as determined by treatment regimens and their department specialties in the EHR, and (3) completed at least two symptom assessments within 15 days of CTX initiation (i.e., the start of a new CTX episode), or at 30-, 60-, or 90- days post-initiation.

### Outcomes of interest

We examined all-cause healthcare utilization and death within one year of CTX initiation. Healthcare utilization outcomes included (1) urgent care visits, (2) emergency department (ED) visits, and (3) hospital admissions. Urgent care and ED visits were identified based on department specialties. Our primary outcomes were (1) urgent care visits, (2) ED visits, and (3) death, with focuses on their incidence and time to first occurrence. In secondary analyses, we modeled the incidence rate of hospital admissions, given that about 95% of patients were hospitalized at least once within one year of CTX initiation.

### Symptom assessment using PRO-CTCAE

The PRO-CTCAE evaluated the presence, frequency, severity, and interference of the 12 most common CTX-related symptoms: constipation, decreased appetite, diarrhea, dyspnea, fatigue, fever, insomnia, nausea, paresthesia, pain, rash, and vomiting.^7,24–26,37,38^ Each symptom was assigned a composite score ranging from 0 to 3, with a score of 1 or higher indicating the presence of a symptom.^39,40^

Based on existing evidence^9–11^, the neurological cluster was defined as the number of any fatigue, insomnia, paresthesia, and pain, resulting in a total score ranging from 0 to 4. The GI cluster included constipation, decreased appetite, diarrhea, nausea, and vomiting, yielding a total score ranging from 0 to 5.^11,20–23^

### Demographic and clinical characteristics

Demographic characteristics included age, sex, race/ethnicity, level of education, employment status, and insurance type. Clinical characteristics included cancer site, months since diagnosis, cancer stage, concurrent use of immunotherapy, radiation therapy, and targeted therapy during CTX (all coded as Yes/No), Charlson comorbidity index (CCI), and receipt of specific CTX agents.

### Statistical analysis

We calculated symptom prevalence across four time points. For each symptom, we calculated the prevalence of co-occurring symptoms. Particularly, we were interested in the prevalence of any neurological and GI symptoms, as well as their co-occurring symptoms. We then summarized the most common symptom combinations within each cluster. Finally, we calculated the burdens of neurological and GI clusters over time.

We calculated the incidences of urgent care visits, ED visits, and death within one-year of CTX initiation for the overall population and stratified by age group(< or ≥ 65 years), degree of comorbidity(CCI: 0 or ≥1), time since diagnosis(≤ or > 2 months), cancer stage(≤III or IV), and cancer site(pancreatic cancer or others), as these are important prognostic factors.^41–43^ Time-to-event analysis was used to predict each outcome based on changes in neurological and GI clusters over the first 90 days of CTX. For urgent care and ED visits, we used propensity score weighting to account for immortal time bias introduced by death. We further explored whether the associations were consistent across different prognostic subgroups. We adjusted covariates including age, race, fever, dyspnea, cancer sites, cancer stages, concurrent target therapy, immunotherapy, radiation therapy, time since diagnosis, and comorbidity burden, all of which were associated with healthcare utilization and death.^41–43^ We reported hazard ratios (HR) with 95% confidence intervals (CI).

Secondary analyses were conducted to examine the associations between the burden of symptom clusters within 90 days of CTX and the incidence rate of one-year hospital admission using negative binomial regression. Multiple imputation with 50 replicates was used to handle missing symptom data in all regression analyses.^44^

### Ethics Review

This study was approved by the Dana-Farber/Harvard Cancer Center (DF/HCC) Institutional Review Board (No. 25-048).

## Results

### Overview

Among the 973 patients, more than half were older adults(53%), males(56.7%), White(86.6%), and non-Hispanic(90.5%)(**Table 1**;**Figure 1**). The most common cancer sites were colorectal(39.8%) and pancreatic(27.8%). About 32% of patients had stage IV cancers.

**Figure 1.**
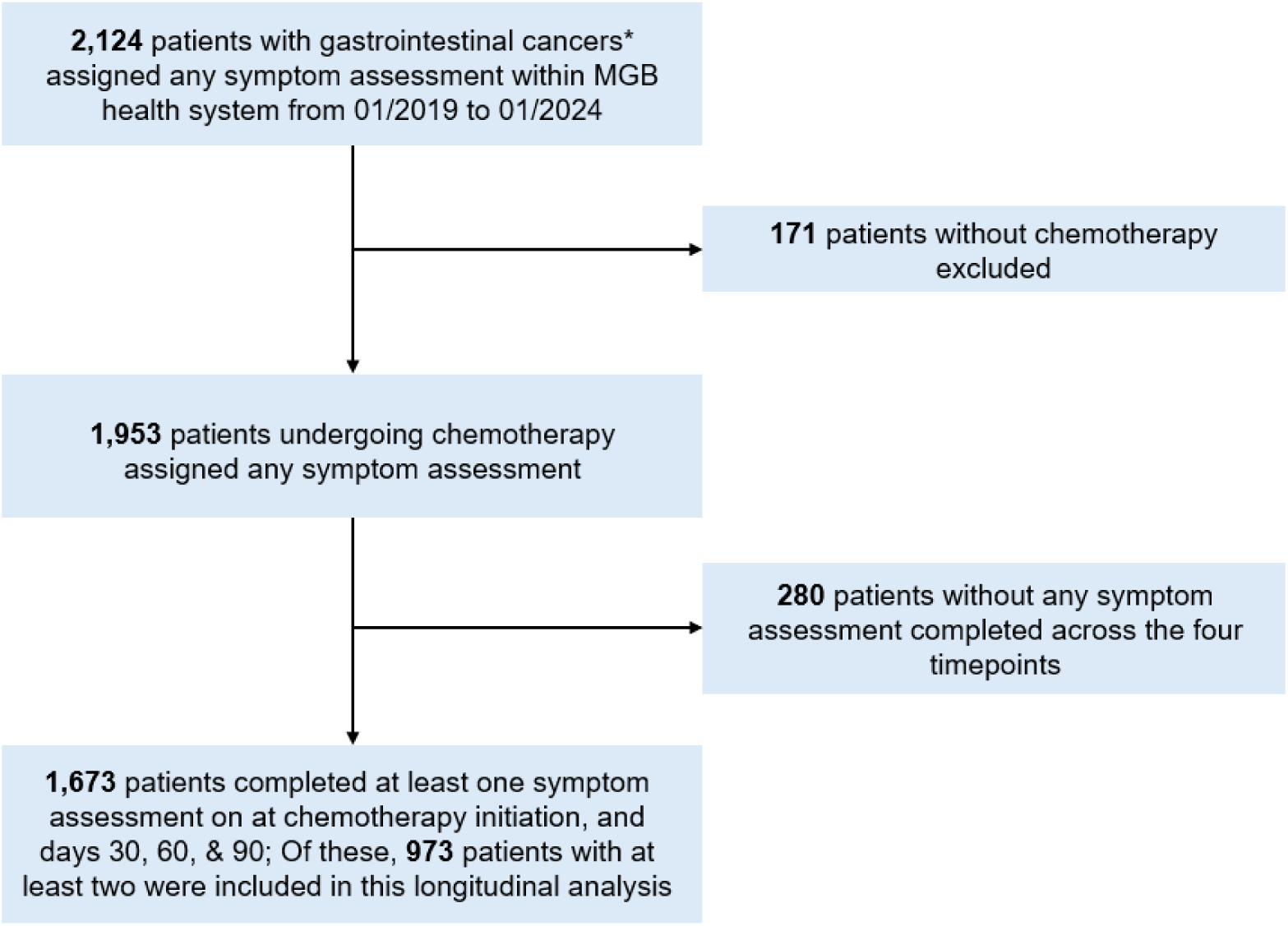
Inclusion and exclusion flowchart for patients with gastrointestinal cancers completed at least two symptom monitoring within the Mass General Brigham Health System, 2019-2024 *: any colorectal, esophagus, liver and bile duct, pancreatic, and stomach cancers

**Table 1.** Demographic and clinical characteristics of patients with gastrointestinal cancers receiving chemotherapy, 2019-2024 (N=973)

| **Characteristics** | **N (%)** |
| --- | --- |
| **≥ 65 years old** | 516 (53) |
| **Male** | 552 (56.7) |
| **White** | 843 (86.6) |
| **Non-Hispanic** | 881 (90.5) |
| **High school or less education** | 261 (26.8) |
| **Unemployed, disabled, retired** | 573 (58.9) |
| **Medicare/Medicaid insurance** | 394 (40.5) |
| **Cancer site** |  |
| Colorectal | 387 (39.8) |
| Esophagus | 136 (14) |
| Liver and bile duct | 127 (13.1) |
| Pancreatic | 270 (27.8) |
| Gastric | 53 (5.5) |
| **Months since diagnosis at chemotherapy initiation (Median, IQR)** | 1.8 (0.8 – 8.4) |
| **Cancer stage** |  |
| 0, I, II, & III | 553 (56.8) |
| IV | 420 (43.2) |
| **Treatment type** |  |
| Concurrent immunotherapy | 241 (24.8) |
| Concurrent radiation | 22 (2.3) |
| Concurrent target therapy | 188 (19.3) |
| **Charleson comorbidity index ≥ 1** | 108 (11.1) |
| **Hospital admission within 90-day from chemotherapy initiation** | 937 (96.3) |
| **Symptom monitoring completion** |  |
| Chemotherapy initiation | 783 (80.5) |
| Day 30 | 833 (85.6) |
| Day 60 | 665 (68.3) |
| Day 90 | 543 (55.8) |

### Symptom prevalence and co-occurrence

At CTX initiation, the most prevalent symptoms were fatigue(72.8%), pain(67.9%), and insomnia(61.1%)(**Table 2**). The most common GI symptoms were decreased appetite(54.7%), diarrhea(50.1%), and constipation(41.5%). More than 75% of patients reported at least two neurological symptoms. Among patients with fatigue, 78.8% experienced pain, and 72.3% reported insomnia(**Figure 2;Table S3**), making “fatigue-insomnia-pain” the most common combination(32%, 188/594;**Table S4**). Over 57.5% of patients had at least two GI symptoms. Among those with decreased appetite, 60.1% had nausea, 59.1% had diarrhea, and 53.7% had constipation(**Figure 2;Table S3**), with the most common GI symptom combination being “constipation-decreased appetite-diarrhea-nausea”(16%, 72/450;**Table S4**). **Table S2** presents symptom prevalence while **Table S3 and Figures S1a - S1c** present the corresponding co-occurring symptoms, from days 30 to 90. Over time, “fatigue-insomnia-paresthesia-pain” and “decreased appetite-diarrhea-nausea” were the two most common combinations.

**Figure 2.**
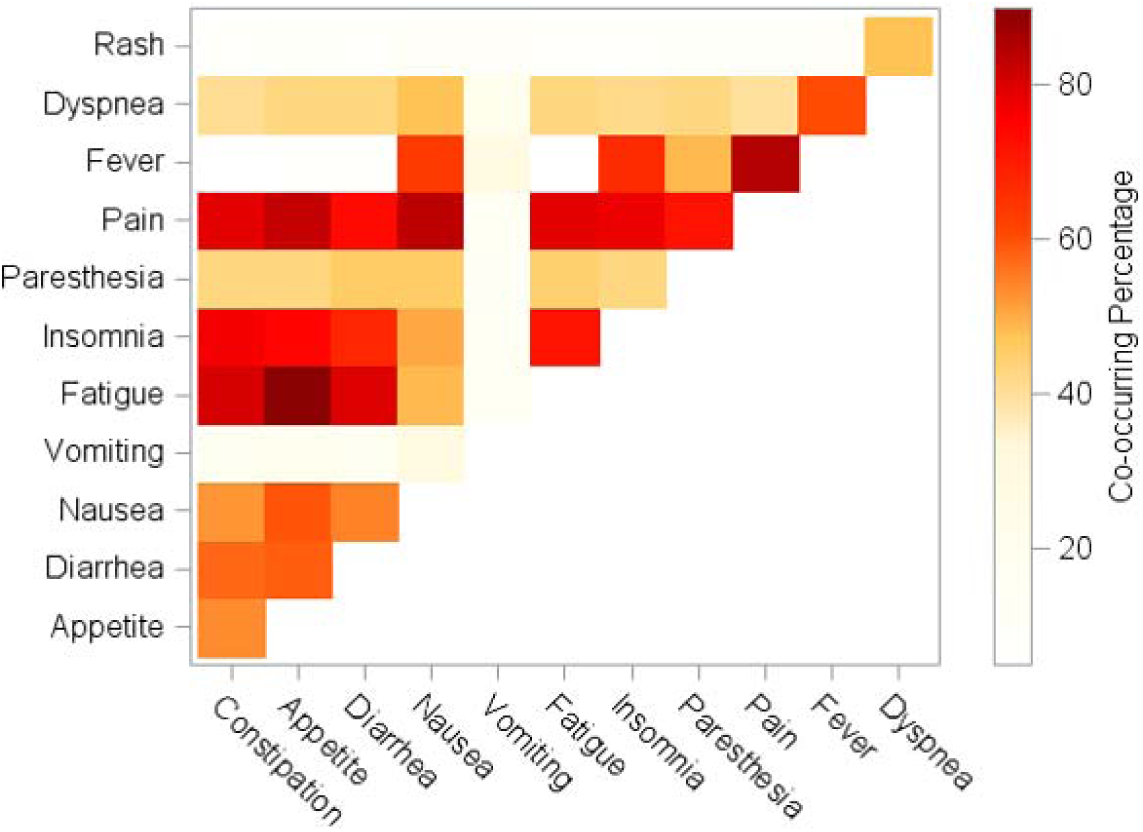
Co-occurring percentages among 12 symptoms in patients with gastrointestinal cancers at chemotherapy initiation (N = 783) *Notes*: 783 out of 973 patients completed symptom monitoring at chemotherapy initiation.

**Table 2.**
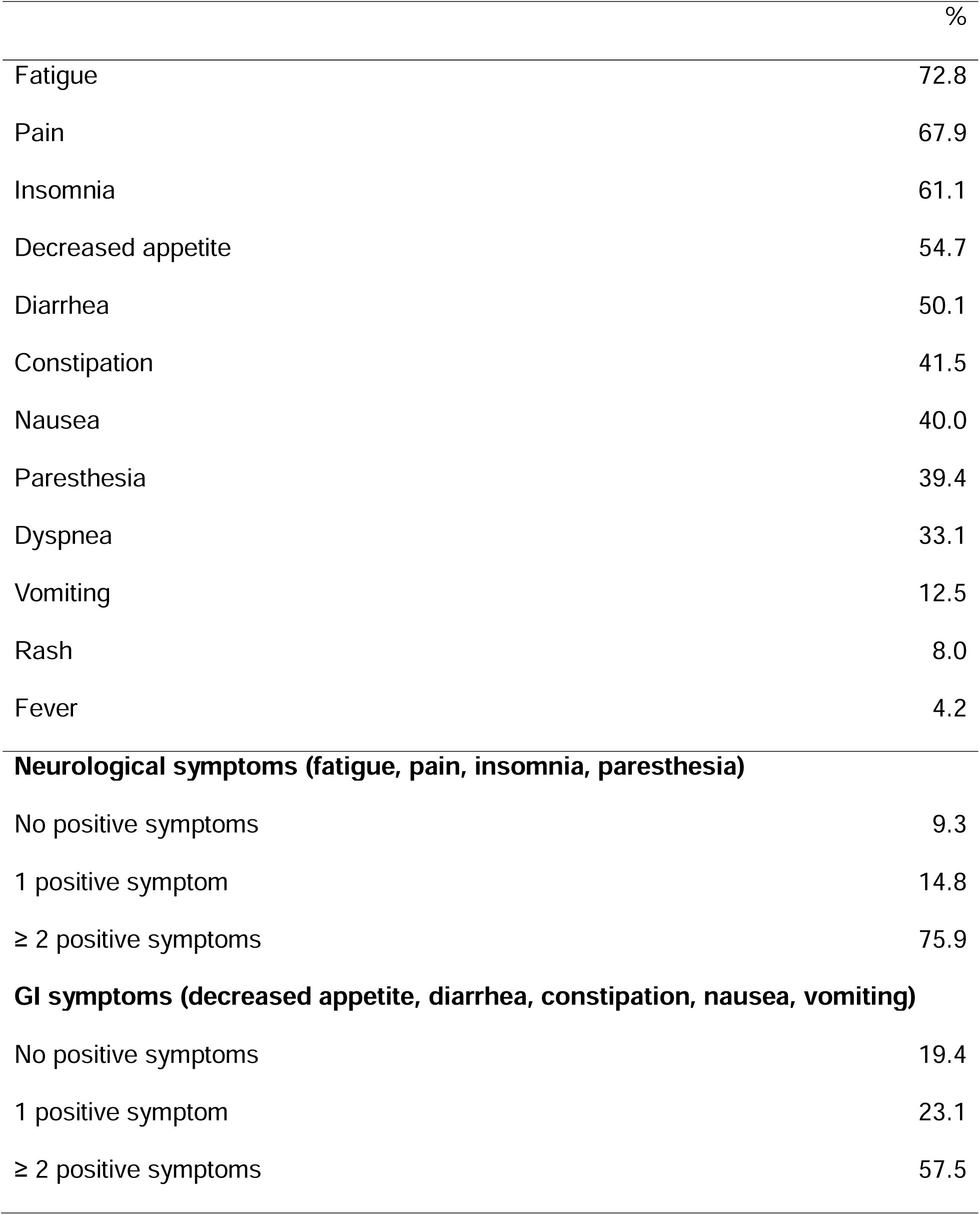
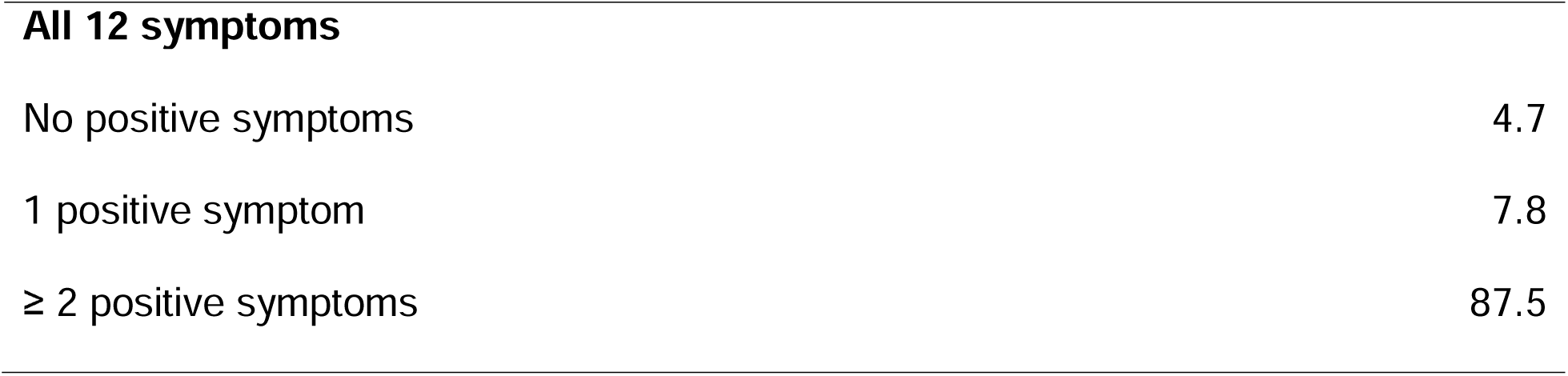
Baseline symptom prevalence of patients with gastrointestinal cancers receiving chemotherapy, 2019-2024 (N = 783)

|  | % |
| --- | --- |
| Fatigue | 72.8 |
| Pain | 67.9 |
| Insomnia | 61.1 |
| Decreased appetite | 54.7 |
| Diarrhea | 50.1 |
| Constipation | 41.5 |
| Nausea | 40.0 |
| Paresthesia | 39.4 |
| Dyspnea | 33.1 |
| Vomiting | 12.5 |
| Rash | 8.0 |
| Fever | 4.2 |
| **Neurological symptoms (fatigue, pain, insomnia, paresthesia)** |  |
| No positive symptoms | 9.3 |
| 1 positive symptom | 14.8 |
| ≥ 2 positive symptoms | 75.9 |
| **GI symptoms (decreased appetite, diarrhea, constipation, nausea, vomiting)** |  |
| No positive symptoms | 19.4 |
| 1 positive symptom | 23.1 |
| ≥ 2 positive symptoms | 57.5 |
| **All 12 symptoms** |  |
| No positive symptoms | 4.7 |
| 1 positive symptom | 7.8 |
| ≥ 2 positive symptoms | 87.5 |

### Burden of neurological and GI clusters over time

Both neurological and GI clusters followed similar patterns over time, showing an increase from days 0 to 30 followed by a gradual decline(**Figure 3**). Throughout the first 90 days of CTX, the burden of the neurological cluster was consistently higher than that of the GI cluster.

**Figure 3.**
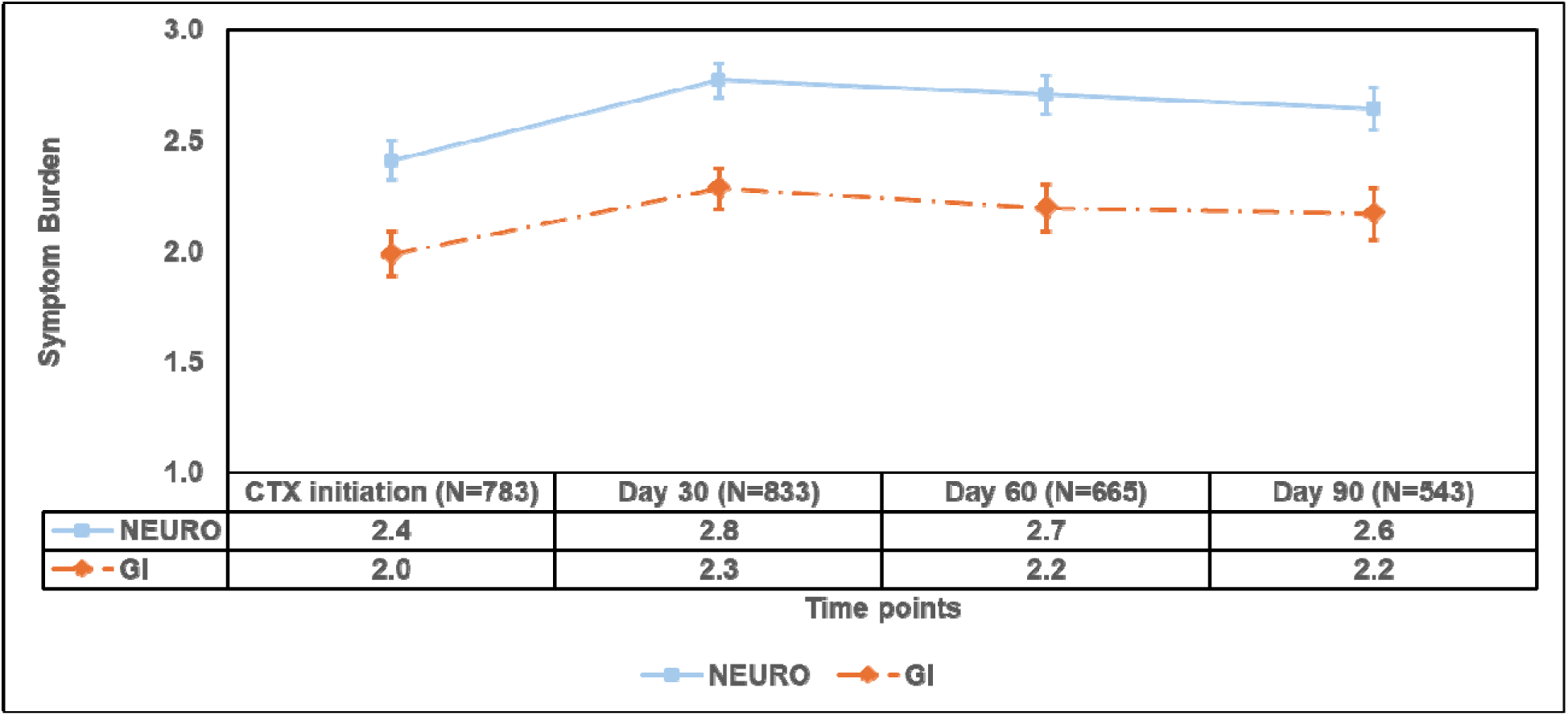
Burden of the neurological and gastrointestinal symptom clusters over time. *Notes:* CTX: Chemotherapy; NEURO: Neurological symptoms (i.e., fatigue, insomnia, paresthesia, and pain); GI: Gastrointestinal symptoms (i.e., constipation, decreased appetite, diarrhea, nausea, and vomiting). Across the four time points, the sample sizes were patients who completed symptom monitoring.

### Healthcare utilization and changes in the burden of neurological and GI clusters

The one-year incidence of urgent care and ED visits were 8.6%(95%CI: 6.9–10.4%) and 53.3%(95%CI: 50.2–56.5%), respectively. A higher incidence of urgent care visits was found among patients with comorbidities, compared to those without(**Figure 4a;Table S5**). Higher incidences of ED visits were found among patients with comorbidities, stage IV cancers, and pancreatic cancer(**Figure 4b;Table S5**). Among patients with urgent care and/or ED visits, there were no significant differences in time to healthcare utilization across levels of burden of neurological and GI clusters at CTX initiation. However, we observed a clear, though non-significant, trend indicating that patients with a higher burden of neurological symptoms at CTX initiation had shorter intervals to their first urgent care visit**(Table S6).** Similarly, patients with a higher burden of GI symptoms at CTX initiation experienced shorter intervals to their first ED visit.

**Figure 4.**
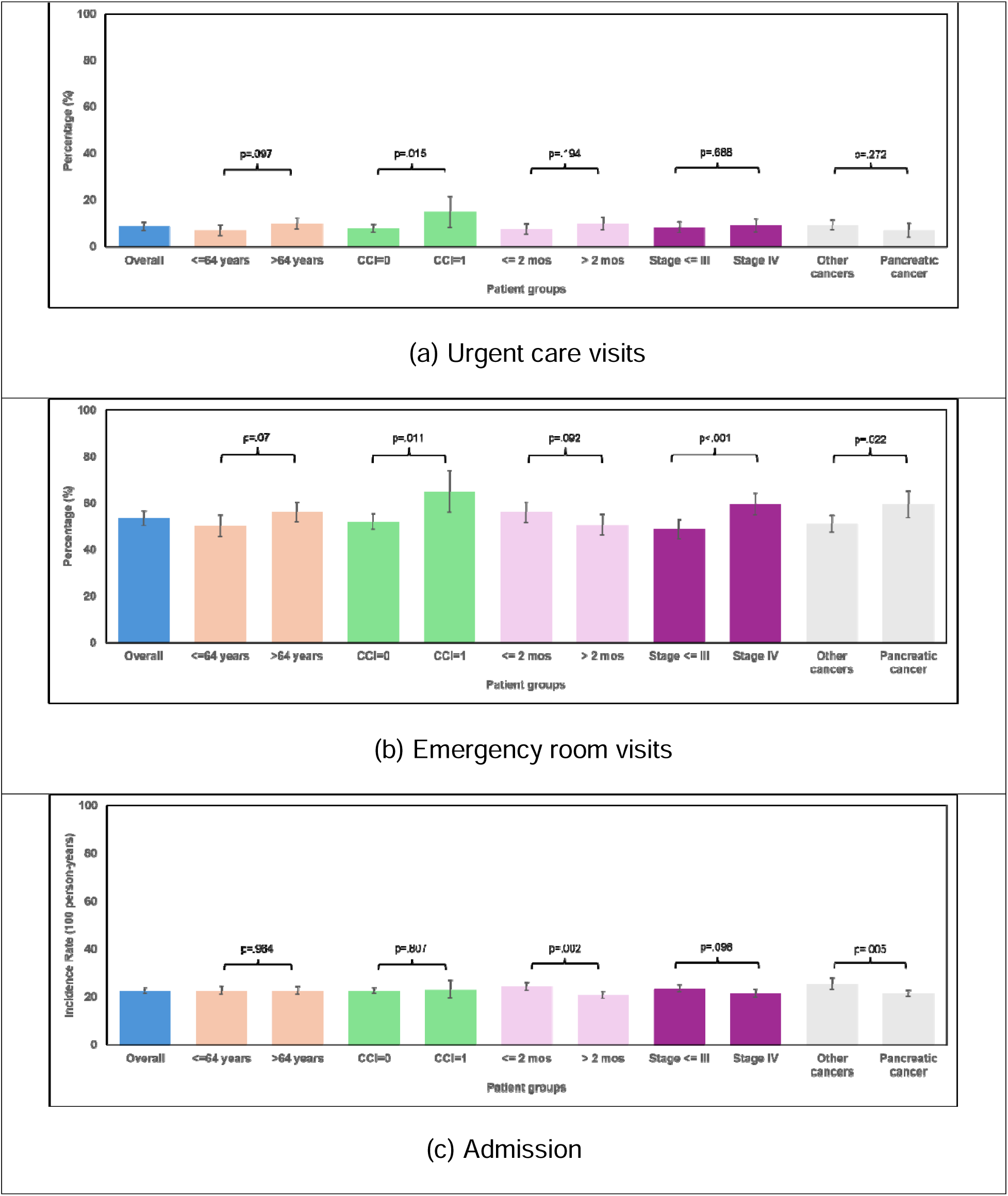
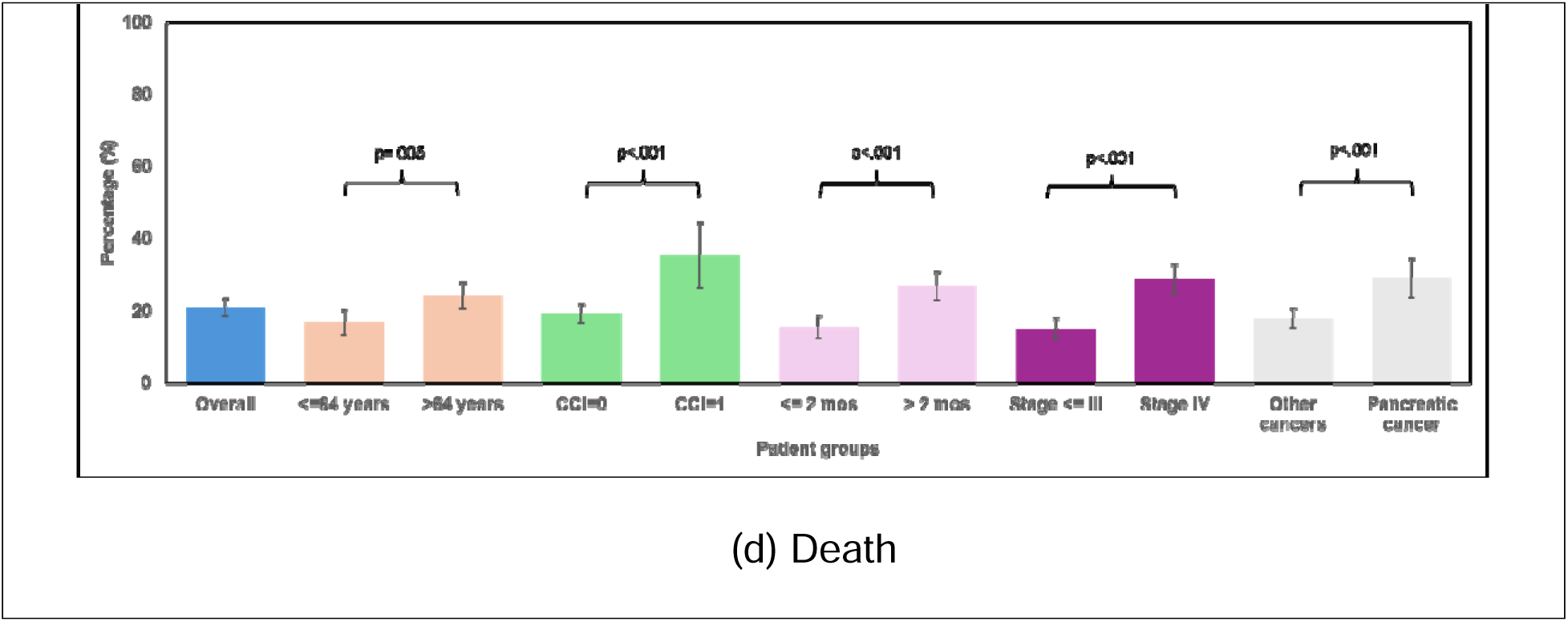
Incidence of outcomes of interests for the overall cohort or stratified by patient subgroups (N=973). *Notes:* CCI: Charlson comorbidity index; mos: Month

The incidence rate of admission was 22.65(95%CI: 21.52–23.84%) per 100 person years, with higher rates found among recently diagnosed patients and those without pancreatic cancer(**Figure 4c;Table S5**).

### Urgent care visits

Time to event analyses found that an increased burden of neurological symptoms from CTX initiation to day 90 post-initiation was associated with a higher risk of urgent care visits(HR: 1.27[95%CI: 0.97–1.67];**Figure 5a**) but not ED visits. Subgroup analyses showed similar trends, with marginally or statistically significant associations observed among older adults(HR: 1.36[95%CI: 0.97–1.90]) and patients without comorbidities(HR: 1.34[95%CI: 1.00–1.80]).

**Figure 5.**
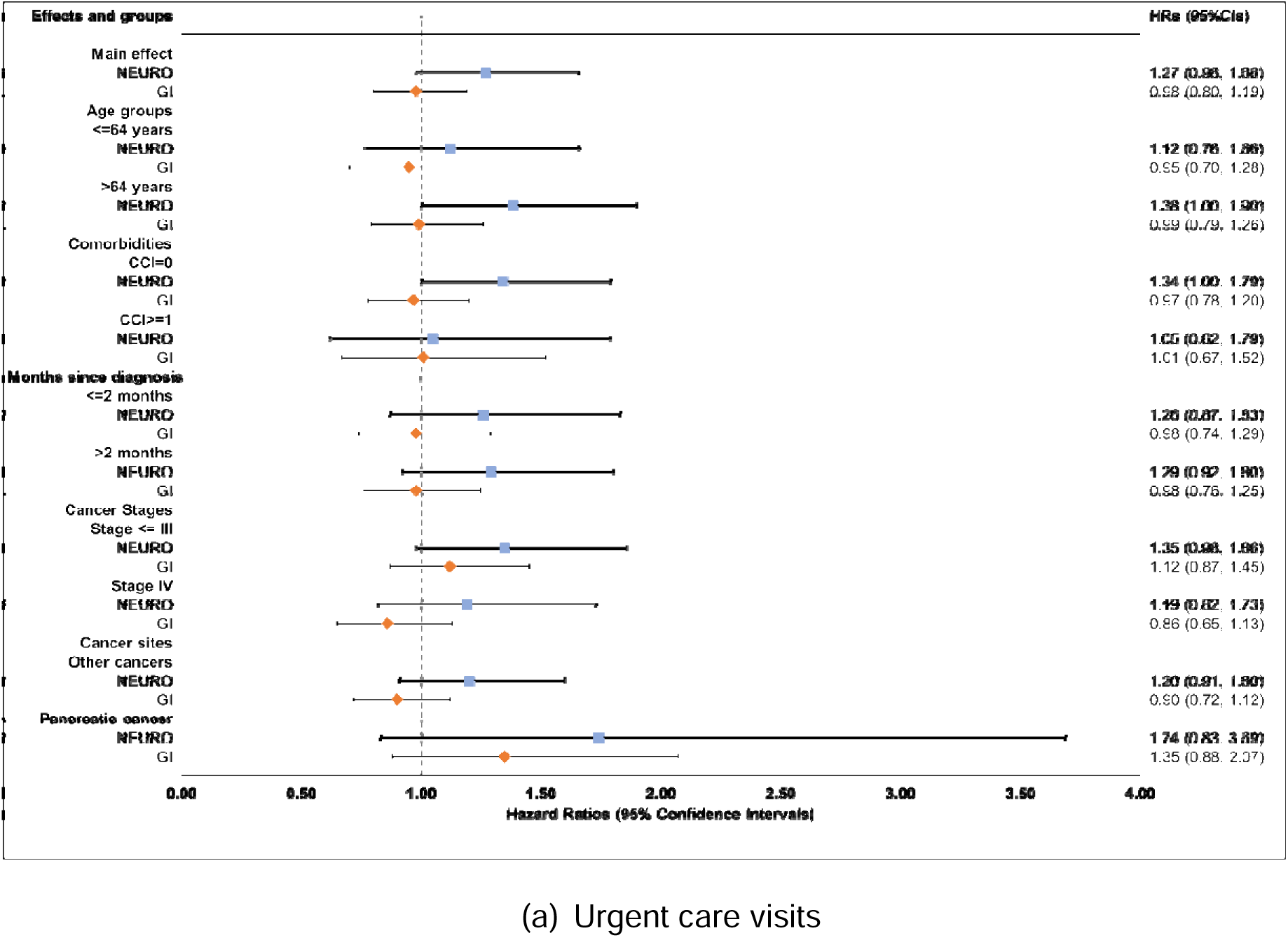
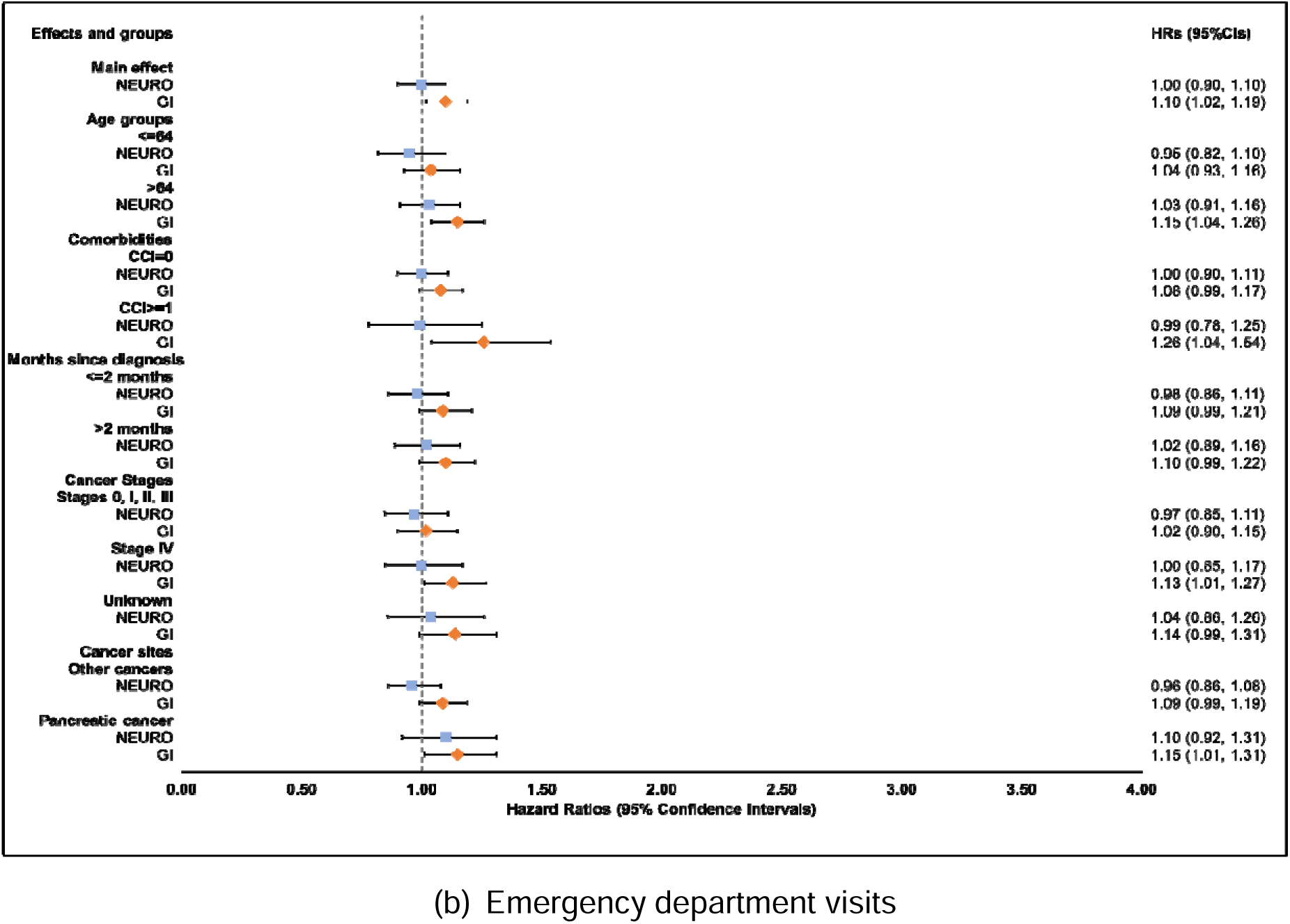
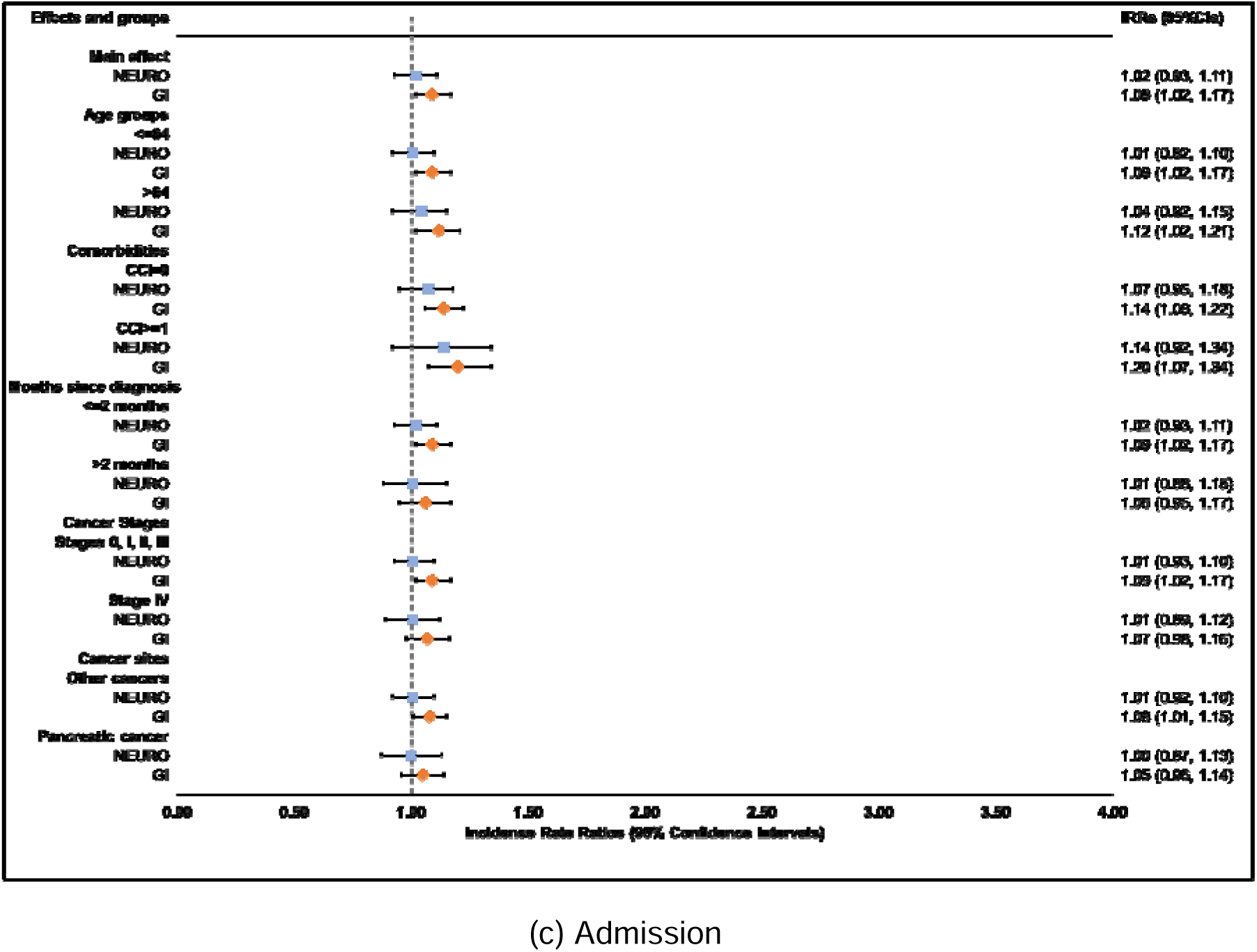
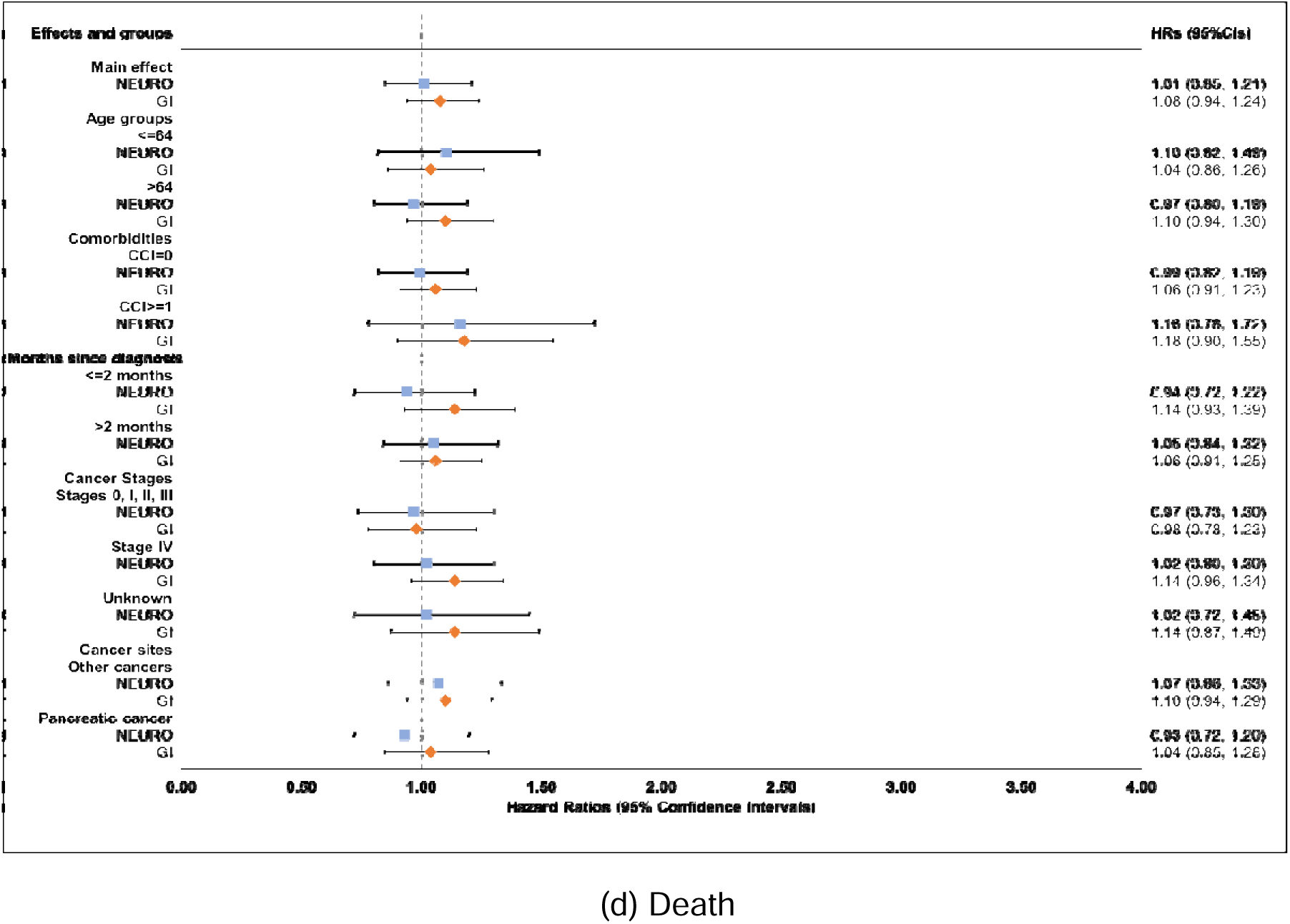
Longitudinal associations of neurological (NEURO) and gastrointestinal (GI) symptom clusters and outcomes of interests (N=973). *Notes:* we adjusted for age, race, cancer sites, cancer stages, fever, dyspnea, concurrent target therapy, concurrent immunotherapy, concurrent radiation therapy, duration after diagnosis, and comorbidities. NEURO: Neurological symptoms (i.e., fatigue, insomnia, paresthesia, and pain); GI: Gastrointestinal symptoms (i.e., constipation, decreased appetite, diarrhea, nausea, and vomiting).

### Emergency department visits

Conversely, an increased burden of GI symptoms from CTX initiation to day 90 post-initiation was associated with a higher risk of ED visits(HR: 1.10[95%CI: 1.02–1.19];**Figure 5b**) but not urgent care visits. Subgroup analyses showed generally consistent trends, with significant associations found among older adults, patients with comorbidities, stage IV cancers, and those with pancreatic cancer.

### Hospital admissions

More than 95% of the patients were hospitalized within 90 days of CTX initiation. The incidence rate of hospital admissions was 22.7(95%CI: 21.5–23.8) per 100 person-years, with significant differences across time since diagnosis and cancer site(**Table S5**). Secondary analyses found that only progression of the GI cluster from CTX initiation to day 90 was significantly associated with the incidence rate of admission(incidence rate ratio [IRR]: 1.09[95%CI: 1.02–1.17];**Figure 5c**), with relatively stronger associations observed among older adults(IRR: 1.12[95%CI: 1.02–1.21) and patients with comorbidities(IRR: 1.20[95%CI: 1.07–1.34]).

### Death and changes in the burden of neurological and GI clusters

The one-year incidence of death was 20.8%(95%CI: 18.2–23.3%). Higher incidences of death were found among older adults, patients with comorbidities, diagnosis more than two months, stage IV cancers, and those with pancreatic cancer(**Figure 4c;Table S5**). There were no significant differences in time to death across levels of burden of neurological and GI clusters at CTX initiation. However, we observed a trend, though non-significant, indicating that patients with a higher burden of GI symptoms at CTX initiation had shorter intervals to death**(Table S6).**

Although not statistically significant, longitudinal regression analysis revealed that the risk of death was positively associated with an increase in GI cluster burden(HR: 1.08[95%CI: 0.94–1.24];**Figure 5d**) but not with the neurological cluster from CTX initiation to day 90 post-initiation. Subgroup analyses revealed suggestive but non-significant associations between the change in GI symptom burden and death among older adults, patients with stage IV cancers, and in those without pancreatic cancer.

## Discussions

Neurological and GI symptoms were confirmed as two prevalent clusters within the first 90 days of CTX initiation. Although the burden of neurological cluster remained higher than that of GI cluster over time, its progression was only associated with an increased risk of one-year urgent care visits, with stronger associations found among older adults and patients without comorbidities. In contrast, progression of the GI cluster was associated with higher risks of one-year ED visits and admission, with stronger effects among older adults and patients with comorbidities.

Our findings reinforce the notion that symptoms often co-occur during CTX. ^5,7,8,19,45^ We defined clusters by the presence of neurological and GI symptoms, as any symptoms detected during CTX should be managed promptly and appropriately to prevent unintended consequences, such as dose reductions and premature treatment discontinuation. The neurological and GI clusters were conceptualized based on commonly used CTX agents, their mechanisms of toxicity, and the corresponding side effects.^9–11,20–32^ Each cluster had consistently prevalent combinations over time, such as “fatigue-insomnia-paresthesia-pain” and “decreased appetite-diarrhea-nausea”.

Neurological and GI clusters showed distinct associations with healthcare utilization. Only the neurological cluster showed a positive association with the risk of urgent care visits. Neurological symptoms are often chronic in nature^46–48^ and may not require emergency-level care, making them more appropriately managed in urgent care centers. The stronger associations found among older adults and/or patients without comorbidities suggest that older patients with lower disease burden may be more likely to visit urgent care centers for managing neurological symptoms. In contrast, GI symptoms tend to be more acute and potentially life-threatening, ^49,50^ often requiring timely medical intervention. This may explain their stronger associations with ED visits and hospital admissions. The stronger effects observed among older adults and patients with comorbidities highlight that, in patients with higher disease burden, managing GI symptoms timely is critical to preventing further deterioration, reducing unplanned ED visits, and even preventing frequent admission.

The GI cluster showed a borderline positive association with mortality, with stronger effects found among older adults, patients with stage IV cancers, and those without pancreatic cancer. No significant association was found between the neurological cluster and mortality. These findings reinforced that GI symptoms and their consequences, such as dehydration and electrolyte imbalance, are generally more acute than neurological symptoms and need timely management.^51,52^ Progressive GI symptoms may increase the risk of death, particularly among older adults with advanced cancers. The borderline association may be attributable to the relatively low incidence of one-year mortality and the small difference in GI symptom burden within the first 90 days of CTX between patients who died and those who survived.

### Implications

Our findings have important implications for enhancing the assessment and management on neurological and GI symptoms to reduce healthcare utilization and mortality. The evidence of co-occurring symptoms underscores the need to raise awareness of these two clusters among both clinicians and patients, enabling more thorough symptom assessments. When clinicians and patients understand which symptoms tend to co-occur, they can better anticipate and explore other potential symptoms when a primary symptom is identified.^12,17,18^ This may uncover symptoms that might otherwise be missed. Moreover, given that GI symptoms may be life-threatening, particularly among older adults with a high disease burden and/or advanced cancers, frequent monitoring of the full range of GI symptoms in high-risk populations may enable earlier interventions and improve treatment outcomes. Symptom co-occurrence also presents opportunities for more cost-effective interventions. For example, by targeting the underlying mechanism for a specific cluster, clinicians may be able to manage multiple symptoms collectively.^12,18,53–55^ Understanding the cascade of symptoms within a cluster and targeting the primary one may also help prevent the development of secondary symptoms.^12,17,18,53,56^ Finally, the distinct associations of neurological and GI symptoms with healthcare utilization suggested that they may require different levels of care. Chronic neurological symptoms, which may require ongoing care, may be more appropriate for urgent care triage, while acute GI symptoms may necessitate ED evaluation and timely management.

### Strengths

Our study utilized a real-world cohort within an integrated health system that has a standardized, well-established PROMs program. We examined whether short-term changes of neurological and GI clusters were associated with long-term healthcare utilization and death, with a rigorous study design to minimize attrition bias and immortal time bias inherent in observational EHR data. We also explored whether these associations were consistent across various prognostic subgroups. These methodological approaches strengthen the internal validity of our findings and support the generation of high-quality evidence with meaningful clinical relevance for routine cancer care.

### Limitations

Although the PRO-CTCAE captured the 12 most common CTX-related symptoms, other symptoms, such as dizziness, heartburn, and bloating, were not included. Additionally, a small number of patients may have received urgent or emergency care outside the MGB system, which could not be captured in this study and may have led to an underestimation of healthcare utilization. What is more, the low incidence of urgent care visits and the small sample sizes of patients with pancreatic cancer or comorbidities may have limited our ability to detect statistical significance. Finally, given that our study was conducted in the Boston metropolitan area, caution is needed when generalizing our findings to other regions of the country.

## Conclusions

Neurological and GI clusters were common among patients with GI cancers receiving CTX. Their progression within the first 90 days from CTX initiation exhibited differential associations with one-year healthcare utilization and death.

## Statement and Declarations

## Acknowledgments

The authors thank the clinicians and program staff for their support on the successful implementation of patient-reported outcome measure collection at the gastrointestinal cancer clinics. We appreciate Mariem Ahmed for helping with manuscript preparation.

## Consent for publication

This is a secondary data analysis based on de-identified electronic medical records. Therefore, no patient consent is needed.

## Data availability statement

Individual patient data will not be shared.

## Ethical approval statement and consent to participate

This study was approved by the Dana-Farber/Harvard Cancer Center (DF/HCC) Institutional Review Board (IRB protocol number: 25-048). This study involves secondary data analysis and does not require patient consent.

## Supplemental Materials

**Table S1.**
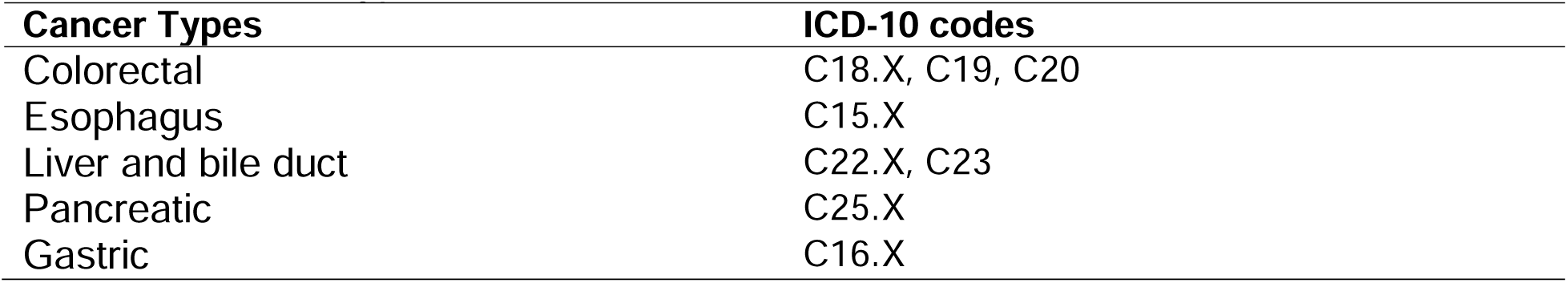
Cancer types and ICD-10 codes.

**Table S2.**
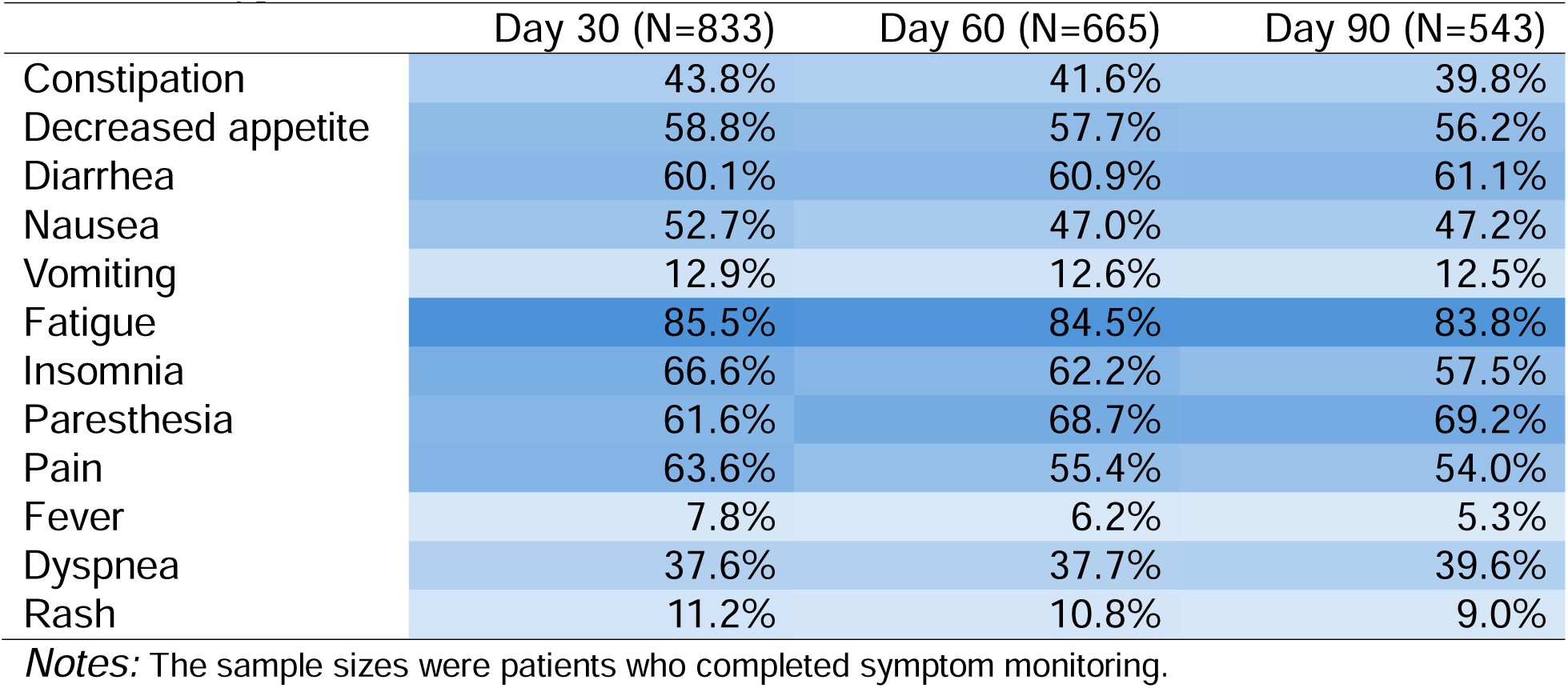
Symptom prevalence of patients with gastrointestinal cancers receiving chemotherapy, 2019-2024.

**Table S3.** Co-occurring symptoms in patients with GI cancers across the four time points of chemotherapy.

| **CTX Initiation**<br>**(N=783)** | Constipation<br>(n=325) | Decreased<br>appetite<br>(n=428) | Diarrhea<br>(n=392) | Nausea<br>(n=313) | Vomiting<br>(n=98) | Fatigue<br>(n=571) | Insomnia<br>(n=479) | Paresthesia<br>(n=308) | Pain<br>(n=532) | Fever (n=33) | Dyspnea<br>(n=259) |
| --- | --- | --- | --- | --- | --- | --- | --- | --- | --- | --- | --- |
| Decreased<br>appetite | 53.7% |  |  |  |  |  |  |  |  |  |  |
| Diarrhea | 57.5% | 59.1% |  |  |  |  |  |  |  |  |  |
| Nausea | 53.2% | 60.1% | 54.6% |  |  |  |  |  |  |  |  |
| Vomiting | 17.5% | 20.1% | 17.6% | 30.40% |  |  |  |  |  |  |  |
| Fatigue | 80.9% | 90.4% | 80.1% | 48.6% | 16.1% |  |  |  |  |  |  |
| Insomnia | 76.6% | 75.0% | 67.9% | 50.8% | 15.7% | 72.3% |  |  |  |  |  |
| Paresthesia | 43.1% | 43.2% | 45.7% | 45.7% | 14.0% | 44.7% | 42.7% |  |  |  |  |
| Pain | 79.4% | 82.9% | 74.0% | 84.0% | 15.6% | 78.8% | 78.2% | 72.1% |  |  |  |
| Fever | 4.9% | 6.3% | 6.4% | 63.6% | 30.3% | 5.4% | 66.7% | 48.5% | 84.9% |  |  |
| Dyspnea | 40.9% | 43.0% | 43.4% | 48.2% | 20.5% | 42.5% | 41.8% | 42.9% | 39.5% | 60.6% |  |
| Rash | 9.2% | 9.8% | 8.7% | 12.1% | 12.7% | 8.6% | 9.4% | 10.1% | 9.6% | 12.1% | 47.6% |
| **Day 30 (N=833)** | Constipation<br>(n=365) | Decreased<br>appetite<br>(n=490) | Diarrhea<br>(n=501) | Nausea<br>(n=439) | Vomiting<br>(n=107) | Fatigue<br>(n=711) | Insomnia<br>(n=555) | Paresthesia<br>(n=513) | Pain<br>(n=530) | Fever (n=65) | Dyspnea<br>(n=313) |
| Decreased<br>appetite | 53.5% |  |  |  |  |  |  |  |  |  |  |
| Diarrhea | 61.6% | 64.5% |  |  |  |  |  |  |  |  |  |
| Nausea | 57.0% | 66.3% | 61.5% |  |  |  |  |  |  |  |  |
| Vomiting | 18.9% | 18.6% | 15.2% | 23.0% |  |  |  |  |  |  |  |
| Fatigue | 90.1% | 95.1% | 88.6% | 56.3% | 13.6% |  |  |  |  |  |  |
| Insomnia | 74.8% | 73.7% | 71.1% | 60.7% | 14.4% | 72.7% |  |  |  |  |  |
| Paresthesia | 66.3% | 62.9% | 66.9% | 65.8% | 13.3% | 64.1% | 66.3% |  |  |  |  |
| Pain | 73.7% | 76.5% | 68.9% | 73.4% | 16.8% | 69.3% | 70.6% | 68.0% |  |  |  |
| Fever | 10.4% | 10.0% | 9.6% | 60.0% | 27.7% | 8.9% | 73.9% | 58.5% | 87.7% |  |  |
| Dyspnea | 46.9% | 46.9% | 42.1% | 44.9% | 17.9% | 42.1% | 44.9% | 40.1% | 45.5% | 56.9% |  |
| Rash | 13.7% | 11.4% | 11.8% | 12.5% | 16.1% | 11.7% | 11.4% | 12.3% | 13.4% | 15.4% | 38.7% |
| **Day 60 (N=666)** | Constipation<br>(n=277) | Decreased<br>appetite<br>(n=384) | Diarrhea<br>(n=405) | Nausea<br>(n=313) | Vomiting<br>(n=84) | Fatigue<br>(n=563) | Insomnia<br>(n=414) | Paresthesia<br>(n=457) | Pain<br>(n=369) | Fever (n=41) | Dyspnea<br>(n=246) |
| Decreased<br>appetite | 49.7% |  |  |  |  |  |  |  |  |  |  |
| Diarrhea | 61.6% | 68.2% |  |  |  |  |  |  |  |  |  |
| Nausea | 56.0% | 59.6% | 55.6% |  |  |  |  |  |  |  |  |
| Vomiting | 16.6% | 19.0% | 16.1% | 24.9% |  |  |  |  |  |  |  |
| Fatigue | 91.0% | 95.1% | 89.1% | 52.0% | 14.0% |  |  |  |  |  |  |
| Insomnia | 74.4% | 73.7% | 67.9% | 56.5% | 15.5% | 70.3% |  |  |  |  |  |
| Paresthesia | 75.0% | 71.5% | 73.8% | 75.3% | 13.8% | 71.4% | 73.9% |  |  |  |  |
| Pain | 65.0% | 68.2% | 59.3% | 68.1% | 18.2% | 60.6% | 66.9% | 57.1% |  |  |  |
| Fever | 8.8% | 9.7% | 7.2% | 78.1% | 34.2% | 7.3% | 82.9% | 78.1% | 95.1% |  |  |
| Dyspnea | 45.3% | 45.2% | 42.5% | 46.2% | 14.6% | 41.8% | 45.5% | 37.9% | 45.9% | 70.7% |  |
| Rash | 11.9% | 10.2% | 10.9% | 10.5% | 9.7% | 10.7% | 10.9% | 11.6% | 12.5% | 12.2% | 37.5% |
| **Day 90 (N=543)** | Constipation<br>(n=216) | Decreased<br>appetite<br>(n=305) | Diarrhea<br>(n=332) | Nausea<br>(n=256) | Vomiting<br>(n=68) | Fatigue<br>(n=455) | Insomnia<br>(n=312) | Paresthesia<br>(n=376) | Pain<br>(n=293) | Fever (n=29) | Dyspnea<br>(n=215) |
| Decreased<br>appetite | 48.9% |  |  |  |  |  |  |  |  |  |  |
| Diarrhea | 66.7% | 69.5% |  |  |  |  |  |  |  |  |  |
| Nausea | 62.5% | 63.6% | 54.8% |  |  |  |  |  |  |  |  |
| Vomiting | 19.4% | 19.7% | 14.2% | 24.2% |  |  |  |  |  |  |  |
| Fatigue | 94.0% | 91.8% | 88.3% | 53.4% | 13.2% |  |  |  |  |  |  |
| Insomnia | 69.4% | 70.8% | 60.5% | 58.3% | 15.1% | 64.0% |  |  |  |  |  |
| Paresthesia | 76.9% | 74.4% | 76.5% | 71.9% | 13.3% | 74.1% | 75.0% |  |  |  |  |
| Pain | 64.8% | 63.6% | 57.2% | 66.8% | 17.1% | 58.0% | 62.2% | 55.6% |  |  |  |
| Fever | 10.2% | 8.2% | 6.9% | 79.3% | 27.6% | 6.4% | 86.2% | 89.7% | 79.3% |  |  |
| Dyspnea | 50.5% | 52.5% | 45.5% | 51.6% | 18.6% | 45.3% | 50.6% | 39.9% | 48.5% | 69.0% |  |
| Rash | 8.8% | 9.8% | 11.8% | 8.6% | 8.2% | 9.2% | 10.6% | 10.4% | 10.9% | 17.2% | 44.9% |

**Table S4.** Prevalent combination of neurological and gastrointestinal symptom clusters over time.

| CTX initiation | Day 30 | Day 60 | Day 90 |
| --- | --- | --- | --- |
| **≥ 2 Neurological symptoms** |  |  |  |
| **n=594** | **n=710** | **n=558** | **n=454** |
| Fatigue-Insomnia-Pain (188 [32%]) | **Fatigue-Insomnia-Paresthesia-Pain (265 [37%])** | **Fatigue-Insomnia-Paresthesia-Pain (198 [35%])** | **Fatigue-Insomnia-Paresthesia-Pain (141 [31%])** |
| **Fatigue-Insomnia-Paresthesia-Pain (154 [26%])** | Fatigue-Insomnia-Pain (112 [16%]) | Fatigue-Insomnia-Paresthesia (95 [17%]) | Fatigue-Insomnia-Paresthesia (84 [19%]) |
| Fatigue-Pain (61 [10%]) | Fatigue-Insomnia-Paresthesia (84 [12%]) | Fatigue-Insomnia-Pain (71 [13%]) | Fatigue-Paresthesia (57 [13%]) |
| Fatigue-Paresthesia-Pain (46 [8%]) | Fatigue-Paresthesia-Pain (68 [10%]) | Fatigue-Paresthesia (62 [11%]) | Fatigue-Paresthesia-Pain (55 [12%]) |
| **≥ 2 Gastrointestinal symptoms** |  |  |  |
| **n=450** | **n=582** | **n=433** | **n=351** |
| Constipation-Appetite-Diarrhea-Nausea (72 [16%]) | **Appetite-Diarrhea-Nausea (91 [16%])** | **Appetite-Diarrhea-Nausea (59 [14%])** | Constipation-Appetite-Diarrhea-Nausea (56 [16%]) |
| **Appetite-Diarrhea-Nausea (48 [11%])** | Constipation-Appetite-Diarrhea-Nausea (76 [13%]) | Constipation-Appetite-Diarrhea-Nausea (58 [13%]) | **Appetite-Diarrhea-Nausea (49 [14%])** |
| Constipation-Appetite (45 [10%]) | Constipation-Appetite-Diarrhea (46 [8%]) | Appetite-Diarrhea (48 [11%]) | Appetite-Diarrhea (41 [12%]) |
| Appetite-Diarrhea (39 [9%]) | Diarrhea-Nausea (46 [8%]) | Constipation-Appetite-Diarrhea (39 [9%]) | Constipation-Appetite-Diarrhea (24 [7%]) |
|  |  |  | Constipation-Appetite-Diarrhea-Nausea-Vomiting (24 [7%]) |

Notes: Percentage was calculated based on the number of patients with at least two neurological or gastrointestinal symptoms at each time point. Clusters in bold consistently had the highest prevalence during follow-up.

**Table S5.**
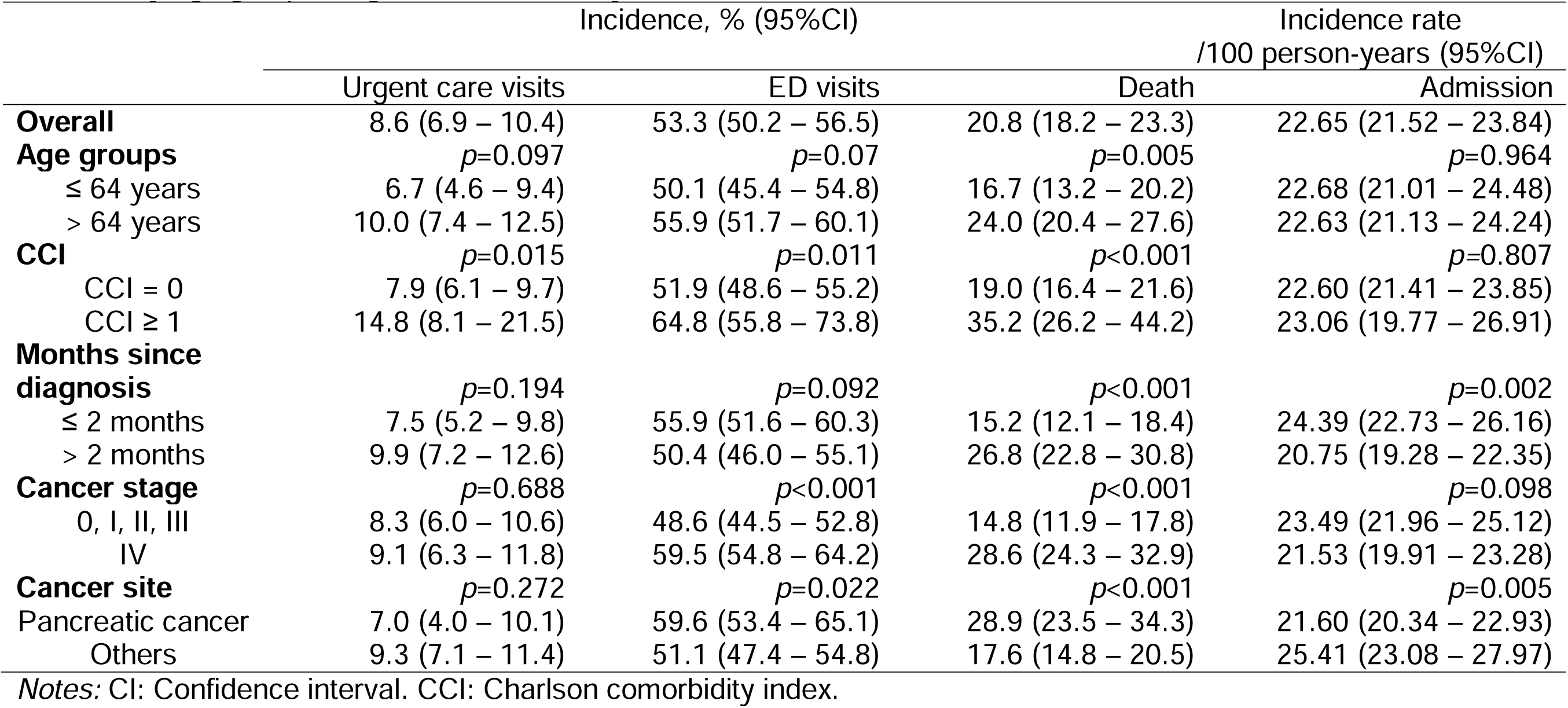
Incidences of one-year urgent care visits, ED visits, death, and admission for the overall population and stratified by age groups, degrees of comorbidity, and cancer sites (N=973)

**Table S6.** Time to first event of one-year urgent care visits, ED visits, and death across burdens of neurological and GI symptom clusters at CTX initiation.

| **Days (Median, IQR)** |  |  |  |  |
| --- | --- | --- | --- | --- |
|  | **Urgent care visits** | **ED visits** | **Death** |  |
| **NEURO** | $p=0.225$ | $p=0.867$ | $p=0.151$ | |
| 0 | 277 (229 – 325) | 98 (22 – 241) | 133.5 (90 – 204) |  |
| 1 | 194 (81 – 230) | 78 (37 – 188) | 208 (132 – 247) |  |
| 2 | 152 (112 – 198) | 87.5 (22 – 181) | 203 (138 – 305) |  |
| ≥3 | 86.5 (54.5 – 221) | 84 (30 – 164) | 156 (101 – 236) |  |
| **GI** | $p=0.924$ | $p=0.644$ | $p=0.174$ | |
| 0 | 127.5 (108 – 174) | 98 (33 – 186) | 203 (95 – 277) |  |
| 1 | 113 (80 – 229) | 81 (22 – 170) | 158 (128 – 239) |  |
| 2 | 141.5 (62.5 – 262.5) | 84 (38 – 198) | 181.5 (125.5 – 300.5) |  |
| ≥3 | 116 (41 – 233) | 75.5 (29 – 163) | 150 (101 – 233) |  |

Notes: CTX: Chemotherapy; IQR: Interquartile ranges; NEURO: Neurological symptoms; GI: Gastrointestinal symptoms.

**Figure S1.**
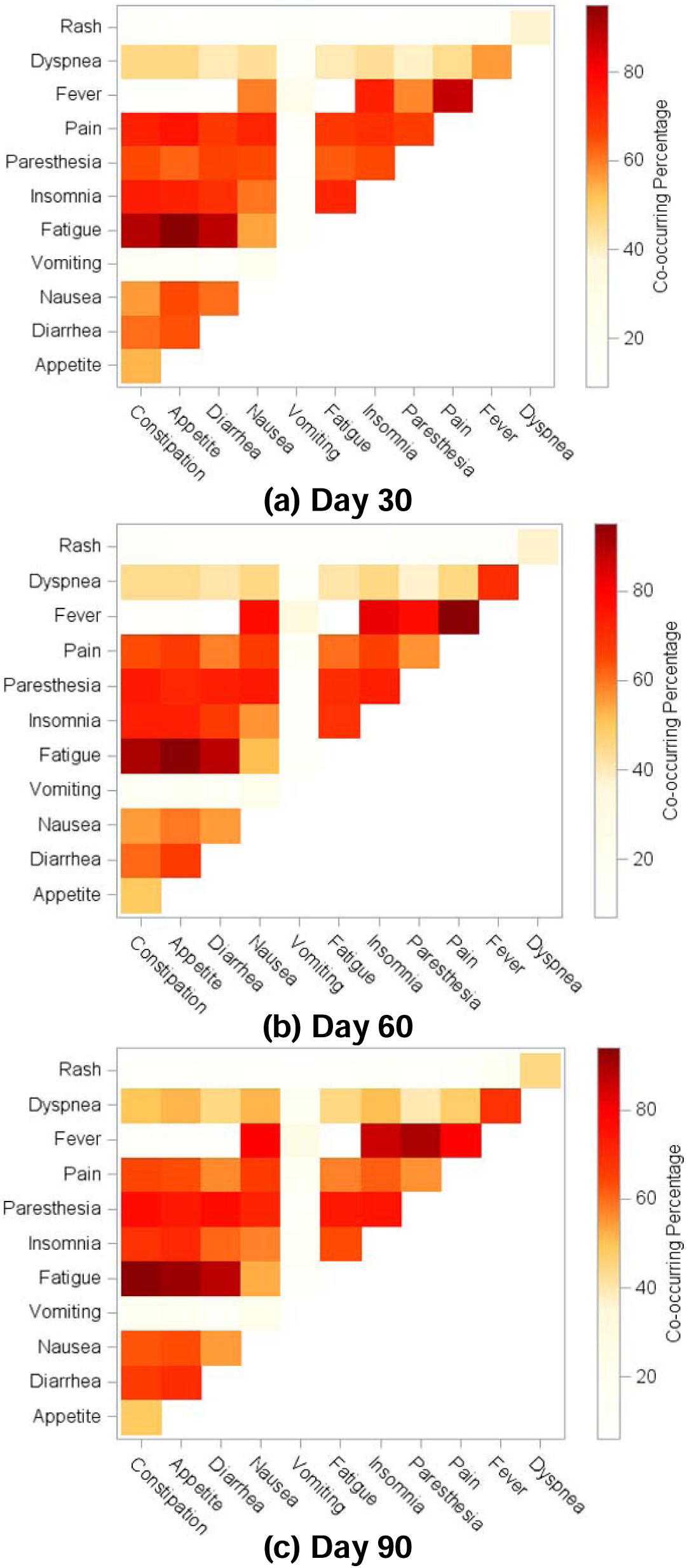
Co-occurring percentages among 12 symptoms in patients with gastrointestinal cancers at days 30, 60 and 90 of chemotherapy initiation (N = 973)

## References

1. U.S. Cancer Statistics Working Group. U.S. Cancer Statistics Data Visualizations Tool, based on 2022 submission data (1999-2020): U.S. Department of Health and Human Services, Centers for Disease Control and Prevention and National Cancer Institute; https://www.cdc.gov/cancer/dataviz, released in November 2023.

2. Barbera L, Atzema C, Sutradhar R, et al. Do patient-reported symptoms predict emergency department visits in cancer patients? A population-based analysis. Ann Emerg Med. Apr 2013;61(4):427–437 e5. doi:10.1016/j.annemergmed.2012.10.010

3. Batra A, Yang L, Boyne DJ, Harper A, Cuthbert CA, Cheung WY. Symptom burden in patients with common cancers near end-of-life and its associations with clinical characteristics: a real-world study. Support Care Cancer. Jun 2021;29(6):3299–3309. doi:10.1007/s00520-020-05827-w

4. Versluis MAJ, Raijmakers NJH, Baars A, et al. Trajectories of health-related quality of life and symptom burden in patients with advanced cancer towards the end of life: Longitudinal results from the eQuiPe study. Cancer. Feb 15 2024;130(4):609–617. doi:10.1002/cncr.35060

5. Han CJ, Reding K, Cooper BA, et al. Stability of Symptom Clusters in Patients With Gastrointestinal Cancers Receiving Chemotherapy. J Pain Symptom Manage. Dec 2019;58(6):989–1001 e10. doi:10.1016/j.jpainsymman.2019.07.029

6. Donlan J, Zeng C, Indriolo T, et al. The Edmonton Symptom Assessment System is a valid, reliable, and responsive tool to assess symptom burden in decompensated cirrhosis. Hepatol Commun. Apr 1 2024;8(4)doi:10.1097/HC9.0000000000000385

7. Harris CS, Kober KM, Conley YP, Dhruva AA, Hammer MJ, Miaskowski CA. Symptom clusters in patients receiving chemotherapy: A systematic review. BMJ Support Palliat Care. Mar 2022;12(1):10–21. doi:10.1136/bmjspcare-2021-003325

8. Han CJ, Reding K, Cooper BA, et al. Symptom Clusters in Patients With Gastrointestinal Cancers Using Different Dimensions of the Symptom Experience. J Pain Symptom Manage. Aug 2019;58(2):224–234. doi:10.1016/j.jpainsymman.2019.04.035

9. Grothey A. Clinical management of oxaliplatin-associated neurotoxicity. Clin Colorectal Cancer. Apr 2005;5 Suppl 1:S38–46. doi:10.3816/ccc.2005.s.006

10. Marupudi NI, Han JE, Li KW, Renard VM, Tyler BM, Brem H. Paclitaxel: a review of adverse toxicities and novel delivery strategies. Expert Opin Drug Saf. Sep 2007;6(5):609–21. doi:10.1517/14740338.6.5.609

11. UpToDate. https://www.uptodate.com/contents/search

12. PDQ Supportive and Palliative Care Editorial Board. Symptom Clusters in Cancer (PDQ®): Health Professional Version. 2024 Jul 11. In: PDQ Cancer Information Summaries [Internet]. Bethesda (MD): National Cancer Institute (US); 2002-. .

13. Miaskowski C, Barsevick A, Berger A, et al. Advancing Symptom Science Through Symptom Cluster Research: Expert Panel Proceedings and Recommendations. J Natl Cancer Inst. Apr 2017;109(4)doi:10.1093/jnci/djw253

14. Dodd M, Janson S, Facione N, et al. Advancing the science of symptom management. J Adv Nurs. Mar 2001;33(5):668–76. doi:10.1046/j.1365-2648.2001.01697.x

15. Miaskowski C, Cooper BA, Melisko M, et al. Disease and treatment characteristics do not predict symptom occurrence profiles in oncology outpatients receiving chemotherapy. Cancer. Aug 1 2014;120(15):2371–8. doi:10.1002/cncr.28699

16. Harris CS, Dodd M, Kober KM, et al. Advances in Conceptual and Methodological Issues in Symptom Cluster Research: A 20-Year Perspective. ANS Adv Nurs Sci. Oct-Dec 01 2022;45(4):309–322. doi:10.1097/ANS.0000000000000423

17. Gutierrez S, Wong R, Milani SA. The pain and depressive symptoms cascade: A bidirectional analysis of the Mexican Health and Aging Study 2012-2015. Int J Geriatr Psychiatry. Oct 2022;37(10)doi:10.1002/gps.5812

18. Kwekkeboom KL. Cancer Symptom Cluster Management. Semin Oncol Nurs. Nov 2016;32(4):373–382. doi:10.1016/j.soncn.2016.08.004

19. Han CJ, Saligan L, Crouch A, et al. Latent class symptom profiles of colorectal cancer survivors with cancer-related cognitive impairment. Support Care Cancer. Sep 5 2023;31(10):559. doi:10.1007/s00520-023-08031-8

20. Yoshisue K, Hironaga K, Yamaguchi S, Yamamoto A, Nagayama S, Kawaguchi Y. Reduction of 5-fluorouracil (5-FU) gastrointestinal (GI) toxicity resulting from the protection of thymidylate synthase (TS) in GI tissue by repeated simultaneous administration of potassium oxonate (Oxo) in rats. Cancer Chemother Pharmacol. 2000;46(1):51–6. doi:10.1007/s002800000123

21. Fleishman SB, Mahajan D, Rosenwald V, Nugent AV, Mirzoyev T. Prevalence of Delayed Nausea and/or Vomiting in Patients Treated With Oxaliplatin-Based Regimens for Colorectal Cancer. J Oncol Pract. May 2012;8(3):136–40. doi:10.1200/JOP.2010.000151

22. Navari RM, Ruddy KJ, LeBlanc TW, et al. Avoidable Acute Care Use Associated with Nausea and Vomiting Among Patients Receiving Highly Emetogenic Chemotherapy or Oxaliplatin. Oncologist. Apr 2021;26(4):325–331. doi:10.1002/onco.13620

23. Xu Y, Villalona-Calero MA. Irinotecan: mechanisms of tumor resistance and novel strategies for modulating its activity. Ann Oncol. Dec 2002;13(12):1841–51. doi:10.1093/annonc/mdf337

24. Andre T, Boni C, Mounedji-Boudiaf L, et al. Oxaliplatin, fluorouracil, and leucovorin as adjuvant treatment for colon cancer. N Engl J Med. Jun 3 2004;350(23):2343–51. doi:10.1056/NEJMoa032709

25. Moore MJ, Goldstein D, Hamm J, et al. Erlotinib plus gemcitabine compared with gemcitabine alone in patients with advanced pancreatic cancer: a phase III trial of the National Cancer Institute of Canada Clinical Trials Group. J Clin Oncol. May 20 2007;25(15):1960–6. doi:10.1200/JCO.2006.07.9525

26. Wang-Gillam A, Li CP, Bodoky G, et al. Nanoliposomal irinotecan with fluorouracil and folinic acid in metastatic pancreatic cancer after previous gemcitabine-based therapy (NAPOLI-1): a global, randomised, open-label, phase 3 trial. Lancet. Feb 6 2016;387(10018):545–557. doi:10.1016/S0140-6736(15)00986-1

27. Hong YS, Nam BH, Kim KP, et al. Oxaliplatin, fluorouracil, and leucovorin versus fluorouracil and leucovorin as adjuvant chemotherapy for locally advanced rectal cancer after preoperative chemoradiotherapy (ADORE): an open-label, multicentre, phase 2, randomised controlled trial. Lancet Oncol. Oct 2014;15(11):1245–53. doi:10.1016/S1470-2045(14)70377-8

28. Conroy T, Hammel P, Hebbar M, et al. FOLFIRINOX or Gemcitabine as Adjuvant Therapy for Pancreatic Cancer. N Engl J Med. Dec 20 2018;379(25):2395–2406. doi:10.1056/NEJMoa1809775

29. Li QJ, He MK, Chen HW, et al. Hepatic Arterial Infusion of Oxaliplatin, Fluorouracil, and Leucovorin Versus Transarterial Chemoembolization for Large Hepatocellular Carcinoma: A Randomized Phase III Trial. J Clin Oncol. Jan 10 2022;40(2):150–160. doi:10.1200/JCO.21.00608

30. Conroy T, Bosset JF, Etienne PL, et al. Neoadjuvant chemotherapy with FOLFIRINOX and preoperative chemoradiotherapy for patients with locally advanced rectal cancer (UNICANCER-PRODIGE 23): a multicentre, randomised, open-label, phase 3 trial. Lancet Oncol. May 2021;22(5):702–715. doi:10.1016/S1470-2045(21)00079-6

31. Xu RH, Muro K, Morita S, et al. Modified XELIRI (capecitabine plus irinotecan) versus FOLFIRI (leucovorin, fluorouracil, and irinotecan), both either with or without bevacizumab, as second-line therapy for metastatic colorectal cancer (AXEPT): a multicentre, open-label, randomised, non-inferiority, phase 3 trial. Lancet Oncol. May 2018;19(5):660–671. doi:10.1016/S1470-2045(18)30140-2

32. Al-Batran SE, Homann N, Pauligk C, et al. Perioperative chemotherapy with fluorouracil plus leucovorin, oxaliplatin, and docetaxel versus fluorouracil or capecitabine plus cisplatin and epirubicin for locally advanced, resectable gastric or gastro-oesophageal junction adenocarcinoma (FLOT4): a randomised, phase 2/3 trial. Lancet. May 11 2019;393(10184):1948–1957. doi:10.1016/S0140-6736(18)32557-1

33. Zeng C, Martin NE, Pusic AL, Edelen MO, Liu JB. Enhancing representativeness of patient-reported outcomes in routine radiation oncology care: a quality improvement protocol to address non-response. BMJ Open. Dec 12 2024;14(12):e097127. doi:10.1136/bmjopen-2024-097127

34. Liu JB, Kaplan RS, Bates DW, Edelen MO, Sisodia RC, Pusic AL. Mass General Brigham’s Patient-Reported Outcomes Measurement System: A Decade of Learnings. NEJM Catalyst. 2024;5(7)doi:10.1056/cat.23.0397

35. Sisodia RC, Dankers C, Orav J, et al. Factors Associated With Increased Collection of Patient-Reported Outcomes Within a Large Health Care System. JAMA Netw Open. Apr 1 2020;3(4):e202764. doi:10.1001/jamanetworkopen.2020.2764

36. Sisodia RC, Rodriguez JA, Sequist TD. Digital disparities: lessons learned from a patient reported outcomes program during the COVID-19 pandemic. J Am Med Inform Assoc. Sep 18 2021;28(10):2265–2268. doi:10.1093/jamia/ocab138

37. Ward Sullivan C, Leutwyler H, Dunn LB, Miaskowski C. A review of the literature on symptom clusters in studies that included oncology patients receiving primary or adjuvant chemotherapy. J Clin Nurs. Feb 2018;27(3-4):516–545. doi:10.1111/jocn.14057

38. Reeve BB, Mitchell SA, Dueck AC, et al. Recommended patient-reported core set of symptoms to measure in adult cancer treatment trials. J Natl Cancer Inst. Jul 2014;106(7)doi:10.1093/jnci/dju129

39. Hinds PS, Pinheiro LC, McFatrich M, et al. Recommended scoring approach for the pediatric patient-reported outcomes version of the Common Terminology Criteria for Adverse Events. Pediatr Blood Cancer. Jun 2022;69(6):e29452. doi:10.1002/pbc.29452

40. Smith AW, Mitchell SA, C KDA, et al. News from the NIH: Person-centered outcomes measurement: NIH-supported measurement systems to evaluate self-assessed health, functional performance, and symptomatic toxicity. Transl Behav Med. Sep 2016;6(3):470–4. doi:10.1007/s13142-015-0345-9

41. Mantziari S, St Amour P, Abboretti F, et al. A Comprehensive Review of Prognostic Factors in Patients with Gastric Adenocarcinoma. Cancers (Basel). Mar 6 2023;15(5)doi:10.3390/cancers15051628

42. Sogaard M, Thomsen RW, Bossen KS, Sorensen HT, Norgaard M. The impact of comorbidity on cancer survival: a review. Clin Epidemiol. Nov 1 2013;5(Suppl 1):3–29. doi:10.2147/CLEP.S47150

43. Gospodarowicz M, O’Sullivan B. Prognostic factors in cancer. Semin Surg Oncol. 2003;21(1):13–8. doi:10.1002/ssu.10016

44. von Hippel PT. How Many Imputations Do You Need? A Two-stage Calculation Using a Quadratic Rule. Sociol Methods Res. Aug 2020;49(3):699–718. doi:10.1177/0049124117747303

45. Li X, Zou Y, Zhang Z, et al. Chemotherapy-related symptom networks in distinct subgroups of Chinese patients with gastric cancer. Asia Pac J Oncol Nurs. Mar 2024;11(3):100366. doi:10.1016/j.apjon.2023.100366

46. Theobald DE. Cancer pain, fatigue, distress, and insomnia in cancer patients. Clin Cornerstone. 2004;6 Suppl 1D:S15–21. doi:10.1016/s1098-3597(05)80003-1

47. Pachman DR, Barton DL, Swetz KM, Loprinzi CL. Troublesome symptoms in cancer survivors: fatigue, insomnia, neuropathy, and pain. J Clin Oncol. Oct 20 2012;30(30):3687–96. doi:10.1200/JCO.2012.41.7238

48. Kroenke K, Lam V, Ruddy KJ, et al. Prevalence, Severity, and Co-Occurrence of SPPADE Symptoms in 31,866 Patients With Cancer. J Pain Symptom Manage. May 2023;65(5):367–377. doi:10.1016/j.jpainsymman.2023.01.020

49. Gallaway MS, Idaikkadar N, Tai E, et al. Emergency department visits among people with cancer: Frequency, symptoms, and characteristics. J Am Coll Emerg Physicians Open. Jun 2021;2(3):e12438. doi:10.1002/emp2.12438

50. Patwari A, Bhatlapenumarthi V, Brann C, et al. Analysis of reasons for Emergency Department (ED) visits and subsequent hospital admissions in patients with solid malignancies: A retrospective study from a cancer center in rural Maine. Journal of Clinical Oncology. 2021;39(28_suppl):241-241. doi:10.1200/JCO.2020.39.28_suppl.241

51. Byar K, Anderson A. An Overview of the Management of Electrolyte Emergencies and Imbalances in Cancer Patients. J Adv Pract Oncol. Mar 16 2025:1–15. doi:10.6004/jadpro.2025.16.7.9

52. Hwa YL, Kull MR. The why and how of maintaining hydration during cancer therapy. Curr Opin Support Palliat Care. Dec 2020;14(4):324–332. doi:10.1097/SPC.0000000000000526

53. Barsevick AM. The elusive concept of the symptom cluster. Oncol Nurs Forum. Sep 2007;34(5):971–80. doi:10.1188/07.ONF.971-980

54. Fleishman SB. Treatment of symptom clusters: pain, depression, and fatigue. J Natl Cancer Inst Monogr. 2004;(32):119–23. doi:10.1093/jncimonographs/lgh028

55. Filetti M, Lombardi P, Giusti R, et al. Efficacy and safety of antiemetic regimens for highly emetogenic chemotherapy-induced nausea and vomiting: A systematic review and network meta-analysis. Cancer Treat Rev. Apr 2023;115:102512. doi:10.1016/j.ctrv.2023.102512

56. Lin SY, Stevens MB. The symptom cluster-based approach to individualize patient-centered treatment for major depression. J Am Board Fam Med. Jan-Feb 2014;27(1):151–9. doi:10.3122/jabfm.2014.01.130145

